# Economic burden of depressive disorders for people living with HIV in Uganda

**DOI:** 10.1101/2024.05.21.24307656

**Authors:** Patrick V. Katana, Ian Ross, Barbra Elsa Kiconco, Patrick Tenywa, Melissa Neuman, Wilber Ssembajjwe, Isaac Sekitoleko, Kenneth Roger Katumba, Eugene Kinyanda, Yoko V. Laurence, Giulia Greco

## Abstract

**Introduction:** Between 8–39 % of people living with HIV (PLWH) in sub-Saharan Africa have depressive disorders (DD). Despite considerable gains in the treatment of PLWH, DD is increasingly recognised as a threat to successful treatment and prevention. PLWH are generally known to suffer from stress and incur higher health-related costs compared to the general population due to care management demand throughout their lifespan. There have been limited studies examining healthcare costs borne by PLWH with DD specifically.

**Objective:** We aimed to estimate the economic burden of DD and HIV amongst PLWH and explore their mechanisms of coping with high out of pocket (OOP) health expenditure.

**Methodology:** This is a cost of illness study nested in an ongoing cluster-randomised trial assessing the effectiveness of integrating treatment of DD into routine HIV care in Uganda (HIV+D trial). The study is using cross-sectional data collected from 1,115 PLWH at trial baseline, using the Patient Health Questionnaire (PHQ-9) to measure DD and a structured cost questionnaire was administered. Forty public health care facilities that provide HIV care in Kalungu, Masaka and Wakiso Districts were randomly selected, and study participants were recruited amongst their patients. Eligibility criteria were patients attending the HIV clinic, aged ≥ 18 years who screen positive for DD (PHQ-9 ≥ 10). Economic costs (OOP expenditure and opportunity costs) were estimated from the household perspective.

**Results:** Mean monthly economic costs amongst those incurring any costs (n=1,115) were UGX 255,910 (US$ 68.64). Mean monthly OOP expenditures were UGX 94,500 (US$ 25.60). On average, respondents missed 6 days of work per month due to healthcare seeking or ill-health for any condition. Key cost drivers were facility bed charges and medication. The majority of respondents (73%) borrowed money from families and friends to cope with the economic burden. About 29.7% reported moderate (PHQ-9 15-19) and 5.12% severe (PHQ-9 ≥ 20) DD symptoms. Respondents with moderate or severe DD had slightly higher average monthly costs than those with mild DD (PHQ-10-14), but the difference was not statistically significant.

**Conclusion:** People living with HIV who experience DD incur in high OOP expenditure and productivity losses. The monthly OOP health expenditure is in the range of 23% of their monthly household income. Social protection mechanisms combined with the integration of the management of DD into routine HIV care could alleviate this burden.

## Background

Globally, approximately 38.4 million people are living with HIV (PLWH) and of these, between 8-39% have depressive disorders (DD) (Kinyanda et al. 2011b; Kinyanda et al. 2017; Myer et al. 2008; Kinyanda et al. 2021). In Uganda, 31% of the estimated 1.4 million PLWH continue to suffer from depression according to a recent meta-analysis (Wagner, Slaughter, and Ghosh-Dastidar 2017). Despite the tremendous progress made to improve access to HIV care in the region, DD remains a significant threat to the fight against HIV, because DD may impact on HIV treatment adherence and risky sexual behaviour. Several studies have shown PLWH suffer from stress and incur higher health-related costs compared to the general population due to care management demands throughout their lifespan (Kinyanda et al. 2011a; Abas et al. 2014; Uthman et al. 2014).

The increased life expectancy for PLWH and DD due to improvements in treatment and care comes with the responsibility of lifelong caring, the cumulative costs for which are substantial (Cohen et al. 2020; Gandhi et al. 2023). This requires patients and family members to bear costs which PLWH and their families are most times not able to bear on their own. These costs can include direct expenses such as medical consultations and drugs (Kirch 2008) or indirect expenses such as reduced productivity (Boccuzzi 2003). Sometimes it can push a family into poverty.

Although antiretroviral therapy (ART) drugs for HIV/AIDS are free of charge for patients at public health centres in Uganda and many parts of the world, patients can still incur costs for comorbid conditions (Pinto et al. 2013). In particular, DD can increase the costs of HIV care through increased visits to health facilities when compared with PLWH without DD (Kinyanda et al. 2017). Most PLWH still incur medical costs (medicine, consultation) to treat opportunistic infections in addition to non-medical costs such as transport and food (Pinto et al. 2013). Furthermore the costs may be catastrophic to people in fragile economic situations in the context of low insurance coverage (Barennes et al. 2015).

To meet the costs of illness, patients and caregivers of PLWH may adopt coping strategies such as the sale of assets or borrowing from relatives or a lending institution (Sauerborn, Adams, and Hien 1996; Russell 1996; McIntyre et al. 2006). Such strategies can help to cope with immediate economic burden; however, they can also be potentially ‘risky’ for their future financial wellbeing. The selling of livelihood assets, for instance, reduces the caregiver’s or family’s ability to generate future income (Flores et al. 2008).

Although several studies have focused on the economic burden of HIV on patients’ wellbeing, quality of life and mental health (Maheswaran et al. 2018; Jakubowski et al. 2022; Katana et al. 2020) no studies have been found that evaluate caregivers’ economic burden and its association with severity of DD. It is against this background that a cost of illness assessment within a cluster-randomised trial of integrated management of depression into routine HIV care (HIV+D) was undertaken. Using the patient cost data collected at baseline this study sought to estimate the economic burden of depressive disorders among PLWH in Uganda. We assessed both direct and indirect costs of illness incurred by PLWH and DD. We also elicited coping startegies employed to meet the economic demands of care and treatment. Finally, we assessed the severity of depressive symptoms that may be associated with meeting the demands of care and treatment for patients living with HIV and DD.

## Method

### Study setting

Uganda is a low-income country with a population of 45 million people and a gross national income per capita of US$884 for the yaer 2021 ((World Bank ; Wagner, Slaughter, and Ghosh-Dastidar 2017). It is estimated that 30% of this population lives below the poverty line of USD 1.77 per person each day (UBOS 2021). A strong association between depression and poverty has been shown, and PLWH are at a much higher risk (Cleary et al. 2020; Rutakumwa et al. 2023). In addition, 9.8% of Uganda’ s gross domestic product (GDP) is spent on healthcare but only 1% of this total health expenditure is allocated to mental health (Molodynski, Cusack, and Nixon 2017). Despite access to free HIV care at point of delivery, PLWH spend close to 10% of their household income seeking health care and are at a risk of being pushed further into poverty because of catastrophic health expenditure (Chuma, Gilson, and Molyneux 2007; Kakaire et al. 2016)

The HIV+D trial was carried out in 40 public health facilities (PHFs) that provide HIV care in semi-urban and rural settings of central Uganda. The central region in Uganda has the second highest prevalence rate of HIV nationally, estimated at 7.6% (Ssebunnya et al. 2021). The HIV+D study was a cluster-randomised trial (Kinyanda et al. 2021) that examined effectiveness and cost-effectiveness of integrating the management of depression into routine HIV Care. A minimum of 10 and a maximum of 30 eligible participants were recruited from each of the 40 PHFs in the districts of Kalungu, Masaka and Wakiso. PLWH who were 18 years and above and were on ART for at least 6 months were recruited through random sampling from HIV care clinics.

### Study design

This was a cross-sectional study nested into the HIV+D trial. Cost data was collected from patients at baseline, 3 months, and 12 months through face-to-face interviews at the PHFs between May 2021 and December 2022. For this study, we used baseline data from the study population of 1,115 PLWH and DD across both trial arms.

### Ethics

Ethical approval to conduct this study was obtained from Uganda National Council for Science and Technology (reference number HS645ES), the Uganda Virus Research Institute (UVRI) Research and Ethics Committee (reference number GC/127/20/04/772), and the London School of Hygiene and Tropical Medicine (LSHTM) Ethics Committee (reference number 22,567) (Kinyanda et al. 2021). Informed consent was obtained from all individuals participating in the study.

### Data Collection

We developed a questionnaire that was used to retrospectively collect patient incurred cost data at the baseline of the HIV+D study from May 2021 to December 2022. The patient cost questionnaire was translated from English to Luganda (the predominantly used local language) and then back translated to ensure accuracy and coherence by the HIV+D Health Economics team in consultation with social scientists that were not part of the HIV+D study and a phsychologist who was a member of the HIV+D study team. Both the latter and the former were experienced with translation of research questionnaires. This baseline patient cost questionnaire tool contained questions on direct and indirect costs, coping strategies, visit type (outpatient and inpatient), household income, household expenditure and sociodemographic information, with specific reference to the previous month only so as to minimise recall bias. A paper questionnaire was administered through face-to-face interviews lasting 30-40 minutes. Completed questionnaires were reviewed by the research team for missing information, then data was entered into the study database in OpenClinica for cleaning and analysis.

### Data analysis

We applied a cost of illness approach and included direct and indirect costs.

Direct costs were measured by combining the average costs per patient for out of pocket (OOP) medical and non-medical expenditure for inpatient and outpatient visits. These included adminidtrative costs e.g consultation fees for clinicians, daily hospital charges, user fees at HCFs, HIV and DD diagnostic test fees, the cost of HIV and DD drugs, supplementary foods, and return travel to HCFs for diagnostic and treatment visits. In addition to regular visits to the HIV clinics, some PLWH also visited the HCF for separate depression treatment. In our study, the costs attributable to both HIV and depression visits were combined.

Indirect costs were estimated by capturing the loss of productive working time and calculating wages using the human capital approach. Productivity losses by patients and caregivers or accompanying persons (as reported by patients) was estimated by multiplying the number of days lost due to DD (which included time for seeking or receiving care or periods of inability to work) by a value for time. We assumed an average adult will work for 8 hours in a day, so a day’s wage lost is equivalent to 480 minutes. To apply a common denominator, all income (in cash or in kind) was converted to a daily rate by using the assumptions of 5 working days in a week on average, 20 in a month, and 260 in a year. If the patient earned a wage, their time was valued using the average of their reported income over the past month from the primary job divided by 20 days. If they had secondary job(s), then we summed the reported income across all jobs to obtain the total daily wage of the person over the past month. Average productivity costs were calculated by multiplying that daily wage by the number of days lost as a result of seeking and receiving HIV and depression care.

The average cost of HIV and DD care was computed as the sum of direct and indirect costs over the duration of one month. Our primary result is the average cost where the denominator includes individuals who reported at least one non-zero cost item (i.e. average cost amongst those incurring costs). We also report the average cost where the denominator is the whole sample (i.e. average cost in the study population include zero or missing values). Missing values were interpreted as zero costs. Costs were collected in Uganda shillings and converted to United States Dollars (US$) for analysis at the average exchange rate for 2022 of 0.003 (currencyconveter)

### Coping strategies

Respondents were asked about coping strategies (e.g. borrowing, use of savings, selling household assests), the source of any borrowed resources (e.g. from friends, a bank, or other sources) and any interest charged on loans.

### Depressive disorders (DD)

The 9-item Patient Health Questionnaire (PHQ-9) (Kaggwa et al. 2022) was administered to screen for eligibility in the trial at baseline. The PHQ-9 provides an indicator of depressive severity, with items scored on a 4-point Likert scale from 0 (not at all) to 3 (nearly every day). Total scores are a sum of item scores, so range from 0 to 27 (worst possible). As a measure of severity, PHQ-9 scores are interpreted as 0-4 (minimal DD), 5-9 (minor), 10-14 (mild), 15-19 (moderate) and 20> (severe). In this study, we used a cut-off ≥10 for eligibility, which maximised sensitivity and specificity in a study conducted in Uganda (Akena et al. 2013). To assess the association between cost and PHQ-9 score, we divide the PHQ-9 score into two groups: mild (10-14) and moderate/severe (>=15).

### Analysis

We used descriptive statistics to summarise all socio-demographic, visit type, coping strategies and depressive symptoms variables. Means and medians of continuous data and proportions for categorical data were summarised. We used principal component analysis (PCA) of asset ownership variables to generate a wealth index and quintiles for the whole household sample (Vyas and Kumaranayake 2006). Since the cost data were highly skewed, we used a generalised linear regression model with gamma family and log link to assess the association between cost and PHQ-9 score and accounted for clustering by clinic. Gamma distributions are commonly used to analyse skewed cost data (Veazie et al. 2023). We considered alternative specifications and considered the Bayesian Information Criterion and distribution of residuals to inform model selection.

## Results

A total of 1,118 people living with HIV and depression symptoms were approached to participate in this sub-study. Three were excluded because they were below 18 years old. Consequently, analysis involved 1,115 respondents who completed the structured patient cost questionnaire. Most of the participants were female (77.0%) and from Wakiso district (Table 1). The mean age of participants was 39 years (SD=12.02) and about 39.3% had no formal education. About a half (48%) were married or living with someone and the majority were Christians (85.6%). The mean PHQ-9 score was 13.7 (SD=3.14). About a fifth applied coping strategies to cope with depression and HIV care management.

**Table 1.** Baseline characteristics of study population.

|  | By gender (1,115, %) |  | By PHQ-9 score (1,115, %) |  |
| --- | --- | --- | --- | --- |
| | Female | Male | PHQ-9 10-14 | PHQ-9 $\geq 15$ |
| **Characteristic (n= 1,115)** | 859 (77.0) | 256 (23.0) | 728 (65.3) | 387 (34.7) |
| **Age (years)** |  |  |  |  |
| 18-25 | 120 (14.2) | 20 (7.9) | 88 (12.3) | 52 (13.6) |
| 26-35 | 311 (36.8) | 53 (20.9) | 237 (33.1) | 127 (33.2) |
| 36-45 | 230 (27.2) | 85 (33.6) | 200 (27.9) | 115 (30.1) |
| 46-55 | 121 (14.3) | 66 (26.1) | 125 (17.4) | 62 (16.2) |
| 56+ | 64 (7.6) | 29 (11.5) | 67 (9.3) | 26 (6.8) |
| **Educational attainment** |  |  |  |  |
| No formal education | 349 (40.6) | 89 (34.8) | 283 (38.9) | 155 (40.1) |
| Primary | 371 (43.2) | 109 (42.6) | 324 (44.5) | 156 (40.3) |
| Secondary | 118 (13.7) | 49 (19.1) | 104 (14.3) | 63 (16.3) |
| Tertiary education | 21 (2.4) | 9 (3.5) | 17 (2.3) | 13 (3.4) |
| **Employment status** |  |  |  |  |
| Engaged in an income generating activity in past month | 124 (14.6) | 29 (11.5) | 95 (13.2) | 58 (15.1) |
| Not engaged in an income generating activity | 724 (85.4) | 224 (88.5) | 623 (86.8) | 325 (84.9) |
| **Marital status** |  |  |  |  |
| Single | 85 (9.9) | 39 (15.2) | 72 (9.9) | 52 (13.4) |
| Married | 391 (45.5) | 145 (56.6) | 346 (47.5) | 190 (49.1) |
| Widowed | 122 (14.2) | 12 (4.7) | 101 (13.9) | 33 (8.5) |
| Separated or divorced | 261 (30.4) | 60 (23.4) | 209 (28.7) | 112 (28.9) |
| **Religion** |  |  |  |  |
| Catholic | 392 (45.6) | 141 (55.1) | 352 (48.4) | 181 (46.8) |
| Protestant | 328 (38.2) | 328 (38.2) | 328 (38.2) | 153 (39.5) |
| Muslim | 139 (16.2) | 25 (9.8) | 111 (15.2) | 53 (13.7) |
| **PHQ-9 score (mean/SD)** |  |  |  |  |
| mean | 13.57 | 14.15 | 11.86 | 17.18 |
| (Median, SD) | (13, 3.06) | (14, 3.37) | (12, 1.49) | (16, 2.39) |
| **Used coping strategies** |  |  |  |  |
| Yes | 176 (20.5) | 40 (15.7) | 136 (18.7) | 80 (20.8) |
| No | 682 (79.5) | 214 (84.3) | 591 (81.3) | 305 (79.2) |
| **Socio-economic status** |  |  |  |  |
| Poor | 153 (17.9) | 68 (27.2) | 143 (19.9) | 78 (20.3) |
| poorer | 181 (21.2) | 44 (17.6) | 151 (21.0) | 74 (19.2) |
| Middle | 177 (20.8) | 52 (20.8) | 147 (20.5) | 82 (21.3) |
| Richer | 162 (19.0) | 46 (18.4) | 140 (19.5) | 68 (17.7) |
| Richest | 180 (21.1) | 40 (16.0) | 137 (19.1) | 83 (21.6) |
| **District** |  |  |  |  |
| Kalungu | 143 (16.6) | 37 (14.5) | 128 (17.6) | 52 (13.4) |
| Masaka | 187 (21.8) | 75 (29.3) | 172 (23.6) | 90 (23.3) |
| Wakiso | 529 (61.6) | 144 (56.2) | 428 (58.8) | 245 (63.3) |

Please include footnotes for all tables where you define the acronyms. Including PHQ-9

### Economic costs

The mean economic cost of illness where the denominator is the number of individuals reporting at least one non-zero cost (n=1,115) was UGX 255,910 (US$ 68.64) per month. The median was UGX 109,702 (US$ 29.43) per month with inter-quartile range UGX 53,000 (US$ 14.22) to UGX 256500 (US$ 68.80). Direct cost constituted 59% of the average total cost. The key drivers of direct costs were daily bed hospital charges for inpatients and HIV medication. The mean economic cost of illness where the denominator was the whole sample (n=1115) was UGX 18840 (US$ 5.05) per month. The median was UGX 0 (US$ 0.00) with inter-quartile range [0-1.61].

The average number of days lost was 6 days per month, and average productivity loss due to HIV and depression were UGX 105,216 (US$ 28.22) per month (Table 2).

**Table 2.** Economic cost to patients with HIV and depression.

|  | N(observation)<br>reporting non-zero<br>cost per item | Mean monthly cost per patient<br>amongst those reporting non-zero<br>cost for that item |  | Median USD<br>[IQR] |
| --- | --- | --- | --- | --- |
| **Direct Costs** |  | UGX (SD) | USD (SD) |  |
| **Direct Medical costs** |  |  |  |  |
| Daily bed charges | 28 | 102,929 (200,022) | 27.27 (52.99) | 10.60 [5.56-39.74] |
| Consultation fees | 24 | 14,208 (11,209) | 3.76 (2.97) | 2.65 [1.32-5.30] |
| User fees | 21 | 22,214 (24,323) | 5.89 (6.45) | 2.65 [1.86-7.95] |
| Laboratory fees | 25 | 16,400 (11,662) | 4.35(3.09) | 5.30 [1.33-5.30] |
| Radiology fees | 6 | 40,000 (30,332) | 10.60 (8.04) | 7.95 [5.30-10.60] |
| Other procedures | 13 | 16,039 (18,054) | 4.25 (4.79) | 2.65 [1.33-5.30] |
| HIV drugs | 12 | 40,667 (11,3356) | 10.78 (30.05) | 1.46 [1.19-4.64] |
| Depression drugs | 34 | 26,132 (39,094) | 6.98 (10.45) | 4.01 [1.87-6.68] |
| Other drugs | 33 | 118,79 (11,238) | 3.18 (3.00) | 1.60 [1.34-4.01] |
| Supplements | 22 | 27,818 (46,347) | 7.44 (12.39) | 2.67 [2.14-4.81] |
| Other medical costs | 30 | 22,733 (31,321) | 6.08 (8.37) | 2.87 [1.87-8.02] |
| **Direct Non-Medical costs** |  |  |  |  |
| Travel | 455 | 8,458 (10,691) | 2.26 (2.86) | 1.60 [1.07-2.67] |
| food | 37 | 17,446 (29,424) | 4.66 (7.86) | 1.34 [0.80-5.35] |
| Other non-medical costs | 36 | 19,311 (36,427) | 5.16 (9.74) | 1.20 [0.29-5.35] |
| **Indirect Costs** |  |  |  |  |
| Informal productivity loss & wages | 78 | 105,216 (472,308) | 28.22 (126.69) | 2.95 [0.52- 9.39] |

The mean out-of-pocket costs was UGX 94,500 (US$ 25.60) per month for the non-zero costs, or about 14.5% of estimated average monthly income of households UGX 652,200 (US$ 174.94), and the median was UGX 33,500 (US$ 8.78), or 5.1% of median monthly income. The average indirect cost respresents about 41% of the total cost. Mean total costs were considerably higher than medians, due to outliers with much higher indirect costs.

### Patient cost and depressive disorder

A total of 726 (65.2%), 330 (29.7%) and 57 (5.12%) of patients had mild, moderate and severe depressive symptoms respectively. Table 3 compares costs for those with moderate/severe depression (34.8%) to those with mild symptoms. The aggregate of both direct and indirect cost for PLWH and depression were higher among those with a moderate and severe compared to those with minor and mild. The mean economic cost of illness amongst individuals reporting non-zero costs was UGX 244,679 (US$ 65.59) per month for people with mild depressive symptoms, compared to 257,474 (US$ 69.02) per month for moderate and severe. However, this difference was not statistically significant (p=0.25).

**Table 3.** Patients costs in USD per month and depression outcomes.

| Cost categories | Mild depressive symptoms (PHQ score 10-14) | | | Moderate & Severe depressive symptoms (PHQ score $\geq 15$) | | | p-value* |
| --- | --- | --- | --- | --- | --- | --- | --- |
|  | N(observation) | Mean costs. (SD) | Median costs [IQR] | N(observation) | Mean costs [IQR] | Median costs [IQR] |  |
| **Direct Costs** |  |  |  |  |  |  |  |
| **Direct Medical costs** |  |  |  |  |  |  |  |
| Daily charges | 20 | 18.19 (28.63) | 10.69 (11.76) | 8 | 50.82 (89.05) | 21.38 (34.75) | 0.107 |
| Consultation fees | 13 | 3.47 (2.09) | 2.67 (4.01) | 11 | 4.18 (3.89) | 2.67 (4.01) | 0.572 |
| User fees | 14 | 6.62 (7.70) | 3.74 (8.02) | 7 | 4.58 (3.05) | 2.67 (5.88) | 0.224 |
| Laboratory fees | 15 | 3.74 (2.47) | 2.67 (4.01) | 10 | 5.35 (3.83) | 5.35 (5.35) | 0.203 |
| Radiology fees | 4 | 12.70 (9.61) | 9.36 (12.03) | 2 | 6.68 (1.89) | 6.68 (2.67) | 0.133 |
| Other procedures | 9 | 5.00 (5.62) | 2.67 (4.01) | 4 | 2.67 (2.00) | 2.41 (2.67) | 0.155 |
| HIV drugs | 8 | 2.17 (2.02) | 1.34 (2.27) | 4 | 28.27 (52.45) | 2.81 (54.39) | 0.000 |
| Depression drugs | 22 | 6.88 (7.63) | 4.68 (5.35) | 12 | 7.17 (14.71) | 3.34 (3.61) | 0.945 |
| Other drugs | 21 | 2.88 (2.43) | 1.60 (2.67) | 12 | 3.70 (3.88) | 2.14 (4.41) | 0.441 |
| Supplements | 15 | 7.77 (13.45) | 2.67 (3.21) | 7 | 6.72 (10.68) | 2.67 (3.47) | 0.813 |
| Other medical costs | 20 | 5.21 (6.41) | 2.67 (5.21) | 10 | 7.80 (11.58) | 3.88 (5.35) | 0.464 |
| **Direct Non-Medical costs** |  |  |  |  |  |  |  |
| Travel | 300 | 2.20 (2.10) | 1.60 (1.60) | 155 | 2.38 (3.94) | 1.60 (1.60) | 0.628 |
| food | 29 | 5.42 (8.72) | 2.41 (4.81) | 8 | 1.90 (1.77) | 1.20 (1.87) | 0.035 |
| Other non-medical costs | 27 | 4.13 (8.01) | 1.07 (5.08) | 9 | 8.27 (13.84) | 1.34 (4.81) | 0.291 |
| **Indirect Costs** |  |  |  |  |  |  |  |
| Informal productivity loss & wages | 47 | 16.73 (60.50) | 2.01 (11.57) | 31 | 35.80 (155.98) | 3.35 (8.68) | 0.396 |

USD United States Dollars, SD Standard deviation, IQR Interquartile range

### Coping strategy

Overall, 19% of study participants employed one or more coping strategy (figure 1), the majority (73%) of whom borrowed from family and friends, followed by other sources and least from banks. We observed higher payback amount than the amount borrowed in most participants and majority were yet to complete servicing their borowed amount.

**Figure 1.**
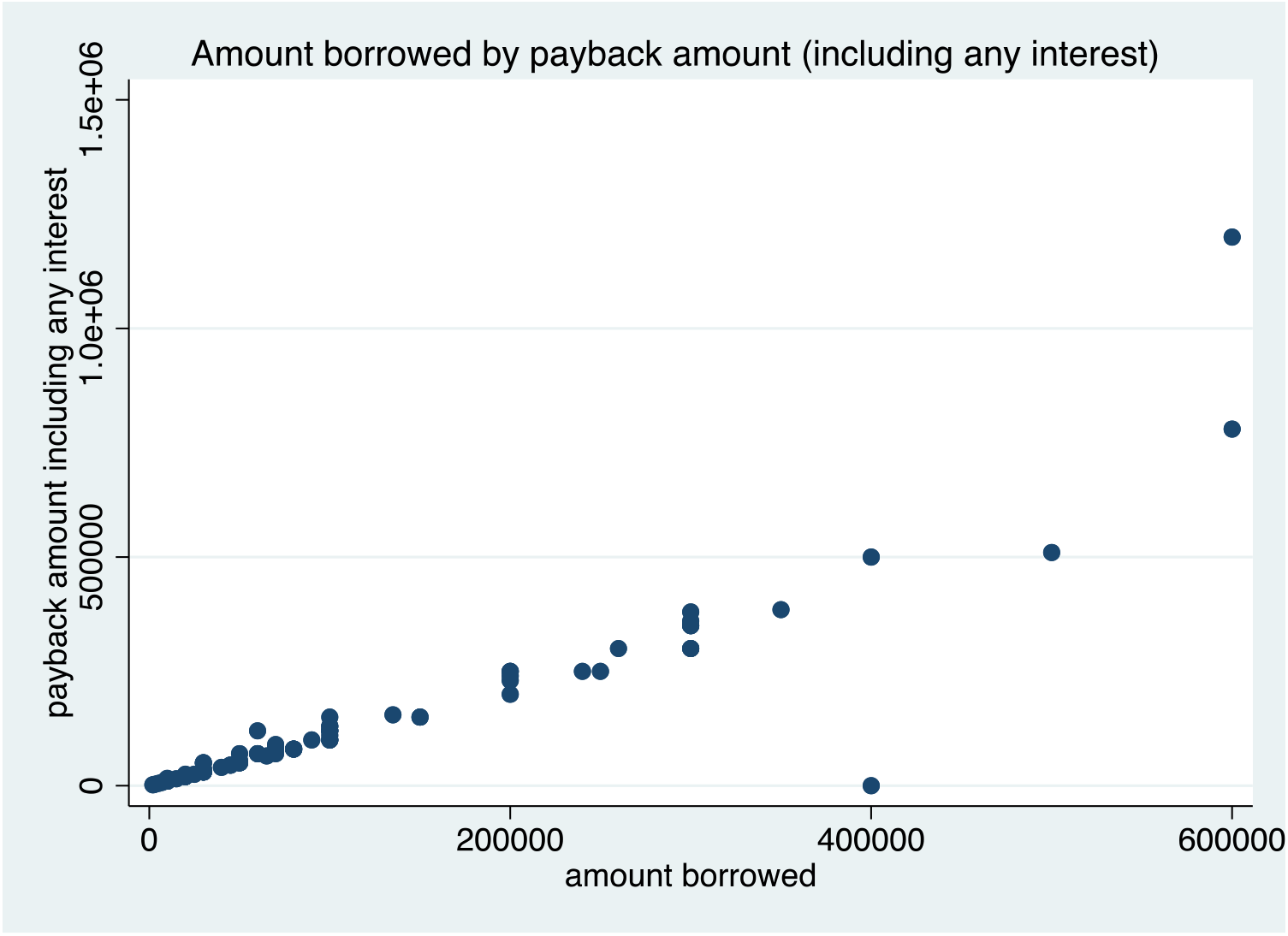
Coping Strategies

## Discussion

This study examined the economic burden of DD for PLWH, using data from 1,115 participants. The majority of our participants were female (77%), which is likely to reflect the higher rates of DD amongst female PLWH in Uganda (Waldron et al. 2021). We observed that patients with moderate and mild depression symptoms had a slightly higher (not statistically significant) economic burden on average compared to those with minor and mild. Amongst those incurring non-zero costs the economic burden was high, representing nearly a quarter of the average monthly household income.

This economic burden is likely to affect individual and household wellbeing (Hughes, Giobbie-Hurder et al. 1999, Lopez-Bastida, Oliva-Moreno et al. 2009, Opoku-Boateng, Kretchy et al. 2017) as financial resources for other basic needs and investments are being diverted to pay for out of pocket health expenditure, and it may result in catastrophic health expenses in the context of fragile economic balance and low insurance coverage (Barennes et al. 2015; Katumba et al. 2023)

The high burden was associated with daily charges for hospitalisation and productivity loss. These results have a two-fold implication on policy planning for health care. First, We hypothesize that the hospitalisation might have resulted from opportunistic diseases hence the need for antiretroviral therapy adherence-enhancing interventions to reduce vulnerability to opportunistic infections (Holmes et al. 2006; Pinto et al. 2013; Katumba et al. 2023). Secondly, productivity loss is an important component often overlooked. Surprisingly few studies have investigated productivity loss in the context of HIV in sub-Saharan Africa. A study in Kenya (Katana et al. 2020) reported lower but comparable productivity costs than our findings. Higher productivity costs might be due to the following reasons: i) HIV and depression care management clinics are located far from the care recipients, ii) poverty among the households which hinders accessibility to healthcare services hence, iii) productivity losses were valued using the average daily reported wage multiplied by the number of days lost while other studies have applied the human capital approach to presentism and absenteeism vi) worse treatment outcomes among the care recipients hence leading to loss of productive time for the patient and the caregiver, and vii) non-zero costs were reported compared to other studies which include zeros for most HIV comprehensive care services.

Patients living with HIV and depression in this setting applied various coping strategies to meet their care needs. We observed that most patients preferred borrowing from friends and relatives. Perhaps this can be explained by the favourable credit terms from friends and family members, unlike banks and other financial institutions, which require collateral and high-interest rates. Some participants borrowed from banks, where they risked losing family assets if they were unable to make payments on time.

Nearly 35% of the patients screened positive for moderate or severe depressive symptoms. Even though not statistically significant, high direct and indirect costs were observed in patients with moderate and severe depressive symptoms. A similar trend was observed in studies by (Katana et al. 2020) and (Harmanci and Cetinkaya Duman 2016).

The study has some limitations; first, the results may be subjected to recall bias by the respondents. Including limiting the recall period to 1 month, and what could have been done, to limit recall bias. Second, we have missing data and zeros, which were excluded from the analysis; as a consequence, these costs may be imprecisely estimated.

Since our study took place at the time of the Covid pandemic there might have been an underestimation of the cost considering the movement restriction during that period; results may to some extent, be generalizable in similar districts in Uganda. Qualitative study on the economic burden of PLWH and depression may help complement our findings. We recommend financial protection/coverage for OOP payments provided for HIV care should be broadened to capture key diseases that are comorbid with HIV (such as DD and TB) so that the outcomes are not worsened. It would also be important to support local community services for DD so that these can be more accessible, particularly for disenfranchised populations.

## Conclusion

The economic burden arising from care and management of HIV and depression remains high and of significant public health concern. Although ARVs are highly subsidized in most HIV clinics in Uganda, patients still incur considerable expenses relative to their incomes. This high economic burden may potentially contribute to depressive symptoms and poor mental health functioning on top of a vicious cycle of poverty. Social programmes to support individuals and families living with HIV is highly recommended.

## Data Availability

The datasets used and/or anlysed in this article are available from the corresponding author on request we will publish the data and analysis codes on OSF.

## Declarations

### Ethics approval and consent to participate

Ethical approval to conduct this was obtained from the Uganda Virus Research Institute (UVRI) Research and Ethics Committee (reference number GC/127/20/04/772), the Uganda National Council for Science and Technology (reference number HS645ES), and the London School of Hygiene and Tropical Medicine (LSHTM) Ethics Committee (reference number 22567). All methods were performed in accordance with relevant local and interntional guidelines and regulations (e.g Declaration of Helsinki). Informed consent was documented in writing for each participant prior to the administration of the questionnaire.

### Consent for publication

Not Applicable

### Availability of data and materials

The datasets used and/or anlysed in this article are available from the corresponding author on request; we will publish the data and analysis codes on OSF.

### Competing interests

The authors declare that they have no competing interests.

### Funding

This study was funded by Wellcome Trust

### Author’s Contributions

YVL, GG & EK conceptualized of the study. YVL, GG designed the study. YVL, GG designed and developed study tools. YVL, GG, PT, WS, BEK, EK & KRK contributed to data collection. PVK, IR, GG & YVL contributed to data analysis. PVK, YVL, GG, MN, & IR contributed to the interpretation of the data. PVK wrote the first draft of the manuscript. YVL, GG, MN, KRK, BEK, PT, IS & IR critically reviewed the manuscript. All authors have read and approved the submitted manuscript.

## Acknowledgements

The authors wish to thank all participants who took part in the study

## Authors information

### Affiliations and Authors

**Department of Global Health and Development, London School of Hygiene and Tropical Medicine, 15-17 Tavistock Place, London, WC1H 9SH, United Kingdom**

Patrick V. Katana, Ian Ross & Giulia Greco

**Department of Disease Control, London School of Hygiene and Tropical Medicine, Keppel Street, London, WC1E 7HT, United Kingdom**

Patrick V. Katana & Ian Ross

**Epidemiology and Population Health and MRC International Statistics and Epidemiology Group, London School of Hygiene and Tropical Medicine**

Melissa Neuman

**MRC/UVRI & LSHTM Uganda Research Unit**

Barbra Elsa Kiconco, Patrick Tenywa, Wilber Ssembajjwe, Isaac Sekitoleko, Kenneth Roger Katumba & Eugene Kinyanda

**King’s College London**

Yoko V. Laurence

## Notes

### Competing Interest Statement

The authors have declared no competing interest.

### Clinical Protocols

https://ijmhs.biomedcentral.com/articles/10.1186/s13033-021-00469-9

### Funding Statement

This study is funded by Wellcome Trust. The funders had no direct role in the design of the study and collection, analysis, and interpretation of data and in writing the manuscript.

### Author Declarations

Ethical approval for this study was obtained from the Uganda Virus Research Institute (UVRI) Research and Ethics Committee (reference number GC/127/20/04/772), the Uganda National Council for Science and Technology (reference number HS645ES), and the London School of Hygiene and Tropical Medicine (LSHTM) Ethics Committee (reference number 22567). Informed consent will be obtained from each participant before enrolment into the study. Further, we got written informed consent from the respondents before conducting the interviews, after informing them about the purpose of the study and their right to withdraw consent at any point. They were also assured of their confidentiality, and that their data would be reported in an aggregated format, and anonymised to protect their identities, throughout the course of this study. I confirm that all necessary patient/participant consent has been obtained and the appropriate institutional forms have been archived, and that any patient/participant/sample identifiers included were not known to anyone (e.g., hospital staff, patients or participants themselves) outside the research group so cannot be used to identify individuals.

